# Investigating the effects of accelerated partner therapy (APT) on chlamydia transmission in Britain: a mathematical modelling study

**DOI:** 10.1101/2020.12.07.20245142

**Authors:** Christian L. Althaus, Catherine H. Mercer, Jackie A. Cassell, Claudia S. Estcourt, Nicola Low

## Abstract

Understanding the effects of partner notification (PN) on the transmission of chlamydia, the most prevalent bacterial sexual transmitted infection worldwide, is critical for implementing optimal control strategies. Accelerated partner therapy (APT) aims to increase the numbers of partners treated and reduce the time to partner treatment. Our objective was to study the effects of APT interventions on partner treatment and chlamydia transmission in order to better understand the results of LUSTRUM, an APT cross-over cluster randomised controlled trial in the UK. We developed a novel deterministic, population-based chlamydia transmission model including the process of PN. We considered a population aged 16–34 years and calibrated the model to sexual behaviour data between people of the opposite-sex and chlamydia prevalence data reported by 3,671 participants in Britain’s third National Survey of Sexual Attitudes and Lifestyles (Natsal-3, 2010–2012) using Approximate Bayesian Computation (ABC). We investigated the potential effects of APT on chlamydia transmission by increasing the number of treated partners and reducing the time to partner treatment compared to standard PN. The median prevalence of chlamydia in the model was 1.84% (95% credible interval, CrI: 1.60%-2.62%) in women and 1.78% (95% CrI: 1.13%-2.14%) in men. Chlamydia positivity was highest in partners of symptomatic index cases with low sexual activity. Infected partners were typically asymptomatic and belonged to the high sexual activity group, i.e., are naturally those infected individuals that will contribute most to onward transmission. Reducing the time to partner treatment without achieving higher numbers of partners treated had only minor effects on reducing chlamydia prevalence. In contrast, the model predicts that a potential increase in the number of partners treated from current levels in Britain (0.51, 95% CrI: 0.21–0.80) by 25% would reduce chlamydia prevalence by 18% (95% CrI: 5%–44%) in both women in men within 5 years. These results suggest that APT, through a potential increase in the proportion of partners treated, would be an effective method to reduce ongoing chlamydia transmission in Britain.

## INTRODUCTION

Partner notification (PN) is considered an important intervention contributing to the control of *Chlamydia trachomatis* (chlamydia) infection. Chlamydia is the most common bacterial sexually transmitted infection (STI) worldwide and primarily found among young sexually active adults (Rowley et al., 2019). Treatment and prevention of chlamydia are of particular importance to women since infection can lead to serious reproductive tract complications (Cates and Wasserheit, 1991). Most infections are asymptomatic and remain undiagnosed, however. PN involves the identification and treatment of sexual partners exposed to infection to prevent re-infection of the index case and prevent onward transmission (Organization and on HIV/AIDS, 1999). The direct effects of PN on the identification of new cases and prevention of re-infection of index cases are well-documented (Golden et al., 2005; Ferreira et al., 2013; Low et al., 2014). The indirect population-level effects of PN on chlamydia transmission, incidence and prevalence are less clear (Golden et al., 2015; Althaus et al., 2012a).

Accelerated partner therapy (APT) is a promising PN strategy, which aims to reduce the time to partner treatment and increase the proportion of partners treated (Estcourt et al., 2012, 2015). APT is an adaption of expedited partner therapy (EPT) (Golden et al., 2015). The difference between the strategies is that APT includes a clinical assessment of sex partners through telephone-led or face-to-face consultation with an appropriately qualified healthcare professional. APT could increase the uptake of PN towards, or even beyond, national targets. Audits of data collected by the National Chlamydia Screening Programme (NCSP) showed that PN uptake (the number of contacts per index case who were reported as having attended a sexual health service within four working weeks of the date of the PN consultation) decreased from 0.53 in 2016 to 0.42 in 2017 and was below the standard of 0.6 (Public Health England, 2018). The national average for the time to attending a health service was 3.2 days (Public Health England, 2016). An exploratory trial of APT resulted in 59% (95% confidence interval, CI: 49%–69%) and 66% (95% CI: 52%–78%) of contactable partners treated in two APT models (telephone hotline and pharmacy), compared to 36% (95% CI: 26%–47%) for standard PN (Estcourt et al., 2012). This suggest that APT has the potential to increase the effective PN uptake. Furthermore, the study found the median time (range) from diagnosis of the index case to partner treatment at 1 (0–14) day and 1 (0–6) day for the two APT models and 4 (0–17) days for standard PN.

Reliable estimates of the effects of APT are needed. The Limiting Undetected Sexually Transmitted infections to RedUce Morbidity (LUSTRUM) programme is a cross-over cluster randomised controlled trial (RCT) in the UK that aims to determine the effectiveness of APT among opposite-sex partners with chlamydia (https://www.lustrum.org.uk). The APT intervention is offered at the level of the sexual health clinic as an additional PN method compared with standard PN (Estcourt et al., 2020). The trial is accompanied by mathematical modelling of chlamydia transmission that we present here in order to better understand the results from the trial and to quantify the effects of APT at the population level. The modelling results will later feed into an economic evaluation for estimating the cost-effectiveness of APT compared to standard PN (Roberts et al., 2012).

Mathematical modelling studies of chlamydia transmission have been widely used to explore and understand the expected population-level effects of screening and PN interventions (Althaus et al., 2012a, 2014; Rönn et al., 2017). Considerable progress in the understanding of chlamydia transmission has been made, but substantial challenges remain. First, it proves difficult to accurately describe the complex sexual contact structure among young adults which is necessary to model PN interventions. Individual-based models offer great flexibility to trace an individual’s partnership history in detail but are difficult to parameterise (Althaus et al., 2012b). In contrast, deterministic, population-based models are much more flexible to fit to epidemiological data, but often restrict PN to ongoing (current) sexual partnerships only (Heijne et al., 2011; Rö nn et al., 2019). Second, the lack of data about chlamydia prevalence in the general population before the introduction of screening for asymptomatic people and PN represents a major source of uncertainty for the parameterisation of mathematical models. For example, a recent modelling study estimated that chlamydia prevalence in the US population would have been almost twice the current levels in the absence of screening and PN (Rö nn et al., 2019). Third, assumptions about the level or duration of immunity following natural clearance of chlamydia has also been identified as an important factor influencing projections of the effects of interventions (Geisler et al., 2013; Omori et al., 2018; Smid et al., 2019).

We take an innovative approach that addresses these earlier limitations in order to adequately quantify the expected effects of different PN strategies on chlamydia transmission. To this end, we developed a novel deterministic, population-based chlamydia transmission model that can be calibrated to multiple data sources within a Bayesian framework. The model includes a dedicated PN module that tracks the most recent partners of index cases and can identify the index-partner combinations that result in the largest effect of PN on reducing chlamydia prevalence. The objectives of this study were to estimate the expected proportions of chlamydia positivity in partners of people with diagnosed chlamydia (index cases) and to quantify the effects of APT on chlamydia prevalence compared with standard PN in Britain.

## METHODS

### Chlamydia transmission model

We developed a deterministic, population-based model of chlamydia transmission between people of the opposite-sex (Fig 1). The model can be described by the following set of ordinary differential equations (ODEs):

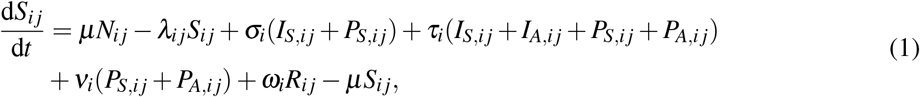

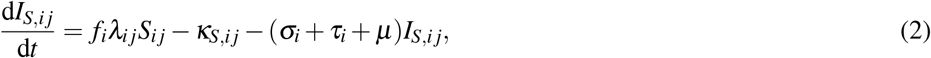

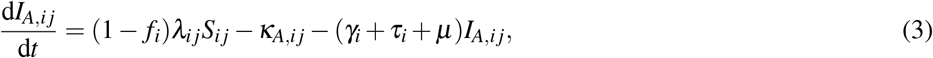

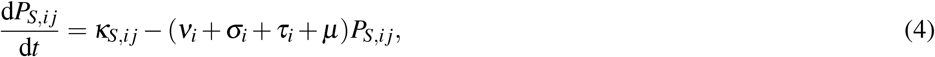

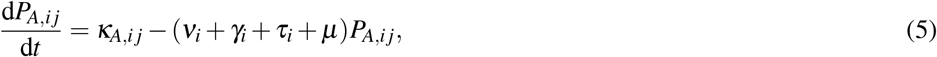

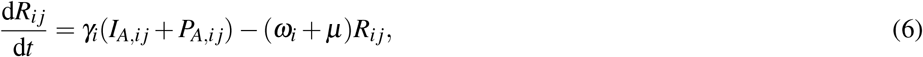

where *S*_*i j*_ are susceptible individuals of sex *i* and sexual activity group *j*, and *N*_*i j*_ denotes the respective population size. A fraction *f*_*i*_ of newly infected individuals is symptomatic (*I*_*S,i j*_) and seeks treatment at rate *σ*_*i*_ or gets tested and treated at rate *τ*_*i*_. Asymptomatically infected individuals (*I*_*A,i j*_) are tested and treated at *τ*_*i*_ or clear the infection spontaneously at rate *γ*_*i*_. Symptomatically and asymptomatically infected individuals can be notified by by chlamydia-positive index cases at rates *κ*_*S,i j*_ and *κ*_*A,i j*_ (see Section *Partner notification*). Notified individuals *P*_*S,i j*_ and *P*_*A,i j*_ are treated at rate *ν*_*i*_. Spontaneous clearance of asymptomatic infection results in the development of immunity (*R*_*i j*_), which is lost at rate *ω*_*i*_. All individuals enter and leave the population at rate *µ*. The force of infection is given by

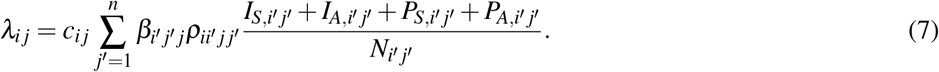

**Figure 1.**
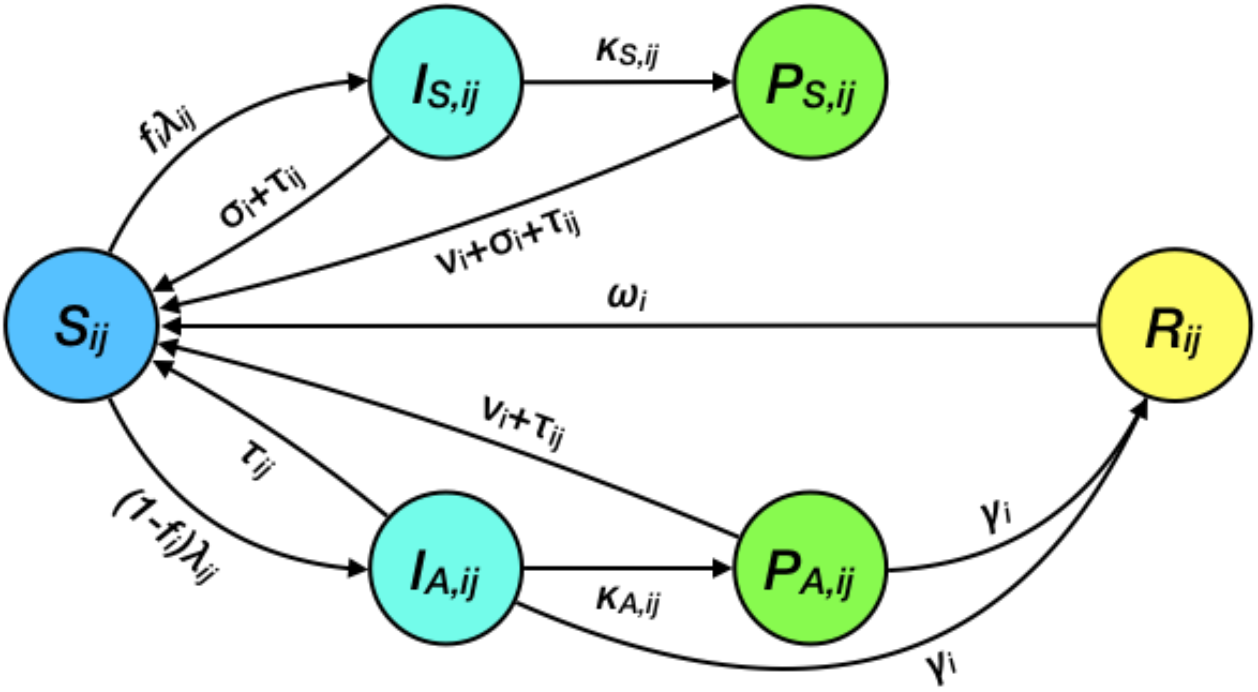
Schematic illustration of chlamydia transmission model. Susceptible individuals *S*_*i j*_ can become symptomatically and asymptomatically infected (*I*_*S,i j*_ and *I*_*A,i j*_). Infected individuals can then become notified (*P*_*S,i j*_ and *P*_*A,i j*_) by their partners. All infected individuals can receive treatment to become susceptible again, or acquire temporal immunity (*R*_*i j*_) through spontaneous clearance of the infection. Movement of individuals into and out of the population is omitted in the scheme. Subscripts *i* and *j* denote sex and sexual activity group, respectively.

Here, *β*_*i*′ *j*′ *j*_ represents the per partnership transmission probability from sex *i′* to *i* and between sexual activity group *j′* and *j*, and *c*_*i j*_ is the sexual partner change rate for individuals of sex *i* and sexual activity group *j. ρ*_*ii*′ *j j*′_ represents the elements of the sexual mixing matrix (Garnett et al., 1999)

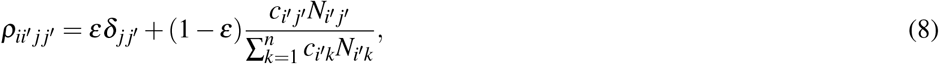

where *δ* _*j j*′_ denotes the Kronecker delta (it is equal to 1 if *j* = *j′* and to 0 otherwise). Sexual mixing between activity groups can vary from proportionate (*ε* = 0) to fully assortative (*ε* = 1).

### Partner notification (PN) module and Who-Notifies-Whom (WNW) matrix

Symptomatically and asymptomatically infected individuals (*I*_*S,i j*_ and *I*_*A,i j*_) can be notified by chlamydia-positive index cases. These index cases who are treated because of symptoms (*σ*) or asymptomatic testing (*τ*) initiate PN of their most recent sexual partner (Fig 2). To describe this process in the transmission model, we constructed a Who-Notifies-Whom (WNM) matrix in PN module. For simplicity, we assumed that PN is offered to index cases only, and not to their notified partners. We further assumed that sexual partners of index cases have not left the population since their last sexual contact. The entries of the WNW matrix can be computed using the following algorithm:

1. Consider index cases (*I*_*S,i*′ *j*′_, *I*_*A,i*′ *j*′_) who can notify their most recent sexual partners that are infected (*I*_*S,i j*_, *I*_*A,i j*_, *I*_*S,n,i j*_, *I*_*A,n,i j*_).
2. Compute the average duration from infection of an index case until diagnosis: 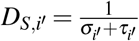 and 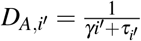.
3. Compute the probability that an index case’s most recent sexual contact was with their infector: 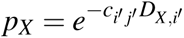, with *X* = {*S,A*}.
  a. Determine the type of infector based on the index case’s force of infection.
  b. Compute the probability that the infection has not yet been cleared in the infector.
4. With probability 1 − *p*_*X*_, the index case’s most recent sexual contact was with an infectee or a other individual.
  a. Determine the type of infectee and other individual based on the index case’s force of infection.
  b. Compute the probability that the infection has not yet been cleared in the infectee or the other individual.

**Figure 2.**
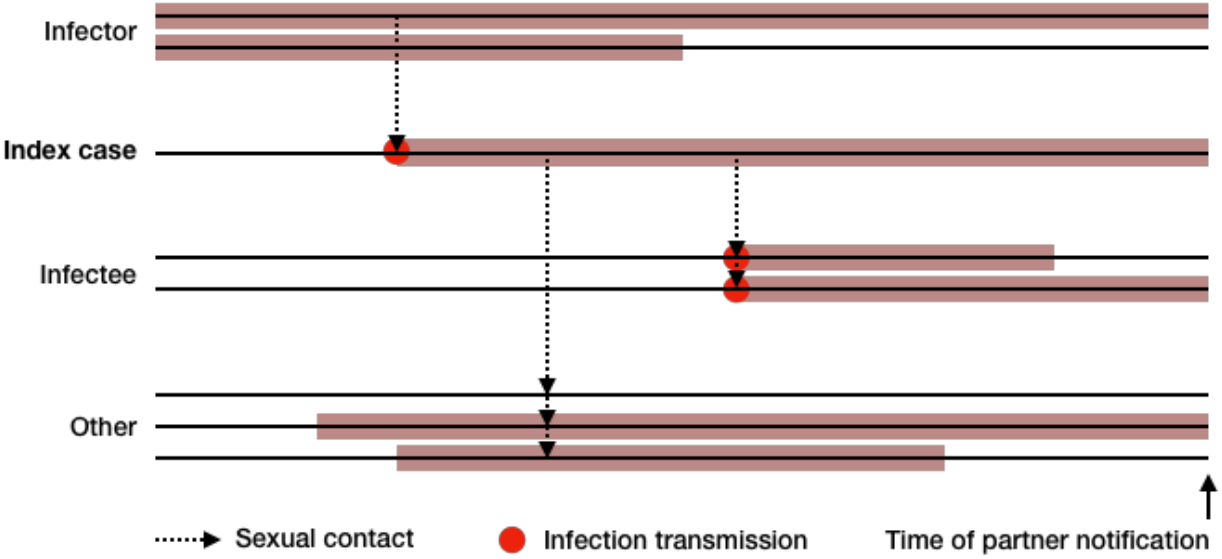
Schematic illustration of partner notification. An index case can either notify their infector (from whom they got infected), an infectee (who got infected by the index case), or another individual (who can either be infected or not). Infector, infectee and other infected individuals can clear the infection before they are notified.

Finally, we multiplied the WNW matrix with the numbers for each index case, the index cases’ rate of diagnoses as defined by *σ*_*i*′_ and *τ*_*i*′_, and the index cases’ proportion of treated partners *f*_*P,i*′_ Summing over all sexual partners of index cases, we obtained the rates *κ*_*S,i j*_ and *κ*_*A,i j*_ at which symptomatically and asymptomatically infected partners of index cases are notified, respectively.

### Data and parameter priors

We estimated sexual behaviour parameters and chlamydia prevalence from Natsal-3, a population-based probability sample survey of sexual attitudes and lifestyles conducted among the resident population in Great Britain from 2010 to 2012 (Erens et al., 2013; Mercer et al., 2013). The full sample consists of 15,162 women and men aged 16–74 years. A subsample of participants aged 16–44 years who reported at least one sexual partner over their lifetime were asked to provide urine samples, resulting in laboratory confirmed chlamydia test results from 2,665 women and 1,885 men (Sonnenberg et al., 2013). We restricted our analysis to 16–34 years old women (*n* = 2,138) and men (*n* = 1,533), which corresponds to the considered age range in the transmission model. We used individual weights to adjust for unequal selection probabilities and to correct for the age, gender and regional profiles in the survey sample. The full datasets of both surveys are available from the UK Data Service Archive at the University of Essex (http://ukdataservice.ac.uk, study number SN7799). The sexual behaviour parameters (*p* and *c*_*i*_ *j*) were estimated from Natsal-3 using a maximum likelihood method as described previously (Althaus et al., 2012a). For the other model parameters, we either used appropriate prior distributions based on literature values or uninformed priors (Table 1).

**Table 1.**
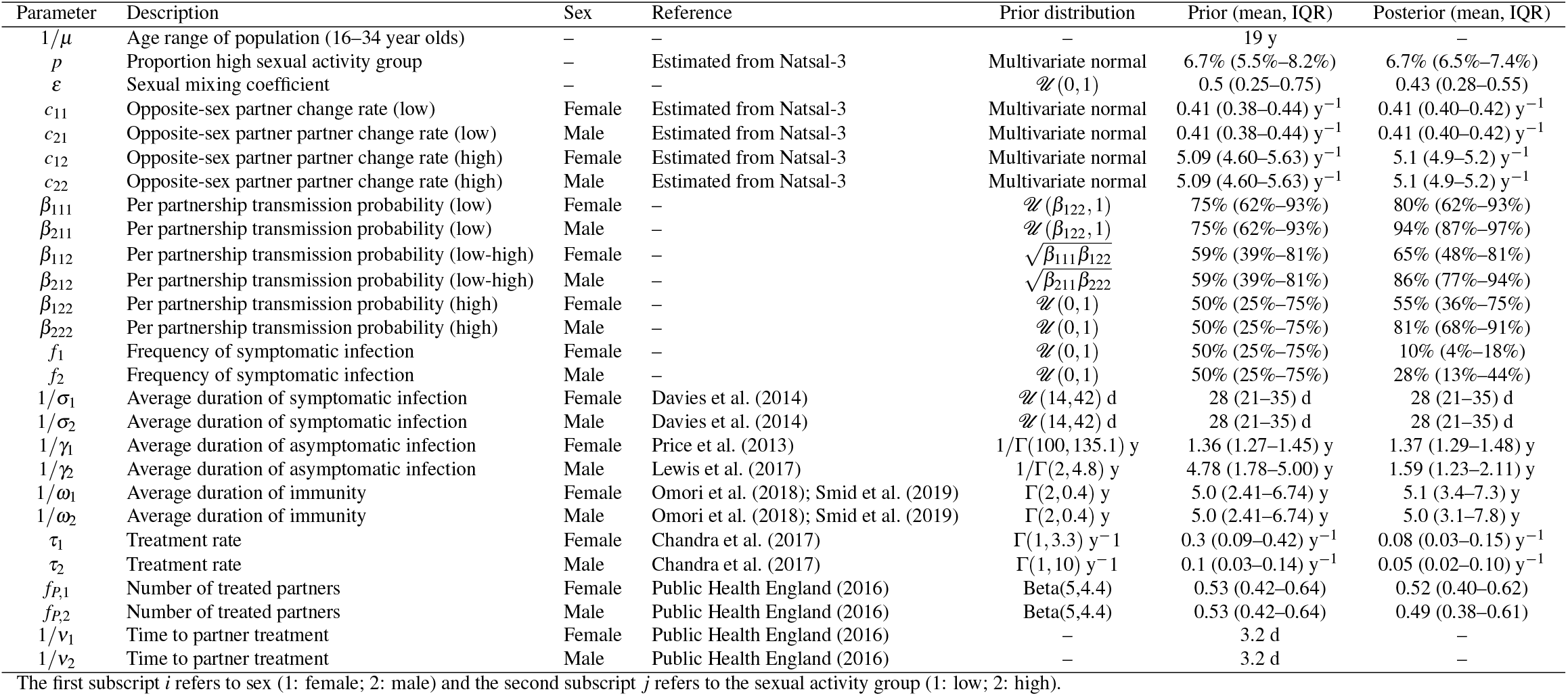
Prior and posterior distributions of model parameters. Parameters *µ* and *ν*_*i*_ were fixed.

### Model calibration, scenarios and APT

We calibrated the model to chlamydia prevalence using Approximate Bayesian Computation (ABC) (Beaumont, 2010). We sampled 10^6^ parameter sets from the prior distributions (Table 1), and used a basic rejection algorithm to select the parameter sets that result in chlamydia prevalences within a specified range (Fingerhuth et al., 2016). We considered the following calibration scenarios:

1. Baseline scenario: Calibration to sex- and activity group-specific prevalence in the presence (current situation) and absence of control interventions.
2. Calibration to sex-specific prevalence in the presence (current situation) and absence of control interventions.
3. Calibration to sex-specific prevalence in the presence of control interventions (current situation).

The estimated chlamydia prevalence from Natsal-3 was 2.2% (95% CI: 1.6%–2.8%) in women and 1.5% (95% CI: 1.0%–2.2%) in men. For the calibration to sex-specific prevalence in presence of control interventions, we accepted those parameter sets where the modelled prevalence was within the 95% CI of these estimates. For the calibration to activity group-specific prevalence in present of control interventions, we assumed that the prevalence in the high activity group cannot exceed 15% in both women and men. Finally, for the calibration to sex-specific prevalence in absence of control interventions, we assumed that the prevalence cannot exceed the point estimate from Natsal-3 by more than twofold (Rönn et al., 2019).

We simulated the effects of APT on chlamydia transmission by 1) increasing the number of treated partners by 5%, 10%, 15%, 20% and 25%, and 2) reducing the time to partner treatment by 1, 2 and 3 days compared to standard PN. We then calculated the relative reduction in prevalence 5 years after the implementation of APT.

We considered scenario 1 as our baseline scenario in the main text and provide all results for scenario 2 and 3 in Supporting Information. Model simulations were performed in the R software environment for statistical computing (R Core Team, 2016) using the function *ode* from the package deSolve (Soetaert et al., 2010). Simulations were run on UBELIX (http://www.id.unibe.ch/hpc), the high-performance computing cluster at the University of Bern. All data and R code files are available on GitHub: https://github.com/calthaus/chlamydia-pn.

## RESULTS

### Model calibration

Calibrating the transmission model to the three scenarios resulted in different posterior distributions of chlamydia prevalence (Figs 3, S1 and S2) and model parameters (Table 1, Figs 4, S3 and S4). In our baseline scenario (scenario 1), the median prevalence of chlamydia in presence of control interventions was 1.84% (95% credible interval, CrI: 1.60%–2.62%) in women and 1.78% (95% CrI: 1.13%–2.14%) in men. The per partnership transmission probabilities were higher for women (male-female transmission) than men (female-male transmission). Sexual mixing between sexual activity groups was weakly assortative (median: 0.43, interquartile range (IQR): 0.28–0.55). 10% (IQR: 4%–18%) of infections were symptomatic in women, compared to 28% (IQR: 13%–44%) in men. The average duration of asymptomatic infection was slightly longer in men (580 d, IQR: 450–470 days) than women (500 d, IQR: 470–540 days). Treatment rates were 0.08 *y*^*−*1^ (IQR: 0.03–0.15 *y*^*−*1^) and 0.05 *y*^*−*1^ (IQR: 0.02–0.10 *y*^*−*1^) for women and men, respectively. For all other parameters, the posterior distributions did not differ substantially from the prior distribution. The alternative calibration scenarios (scenario 2 and 3) where characterised by more assortative mixing, longer durations of asymptomatic infection, and shorter durations of immunity (Figs S3 and S4). Together, this resulted in a somewhat unrealistically high and low chlamydia prevalence in the high and low sexual activity group, respectively (Figs S1 and S2). Hence, we concluded that the baseline scenario (scenario 1) seems most adequate to describe chlamydia transmission between people of the opposite-sex and to quantify the effects of PN.

**Figure 3.**
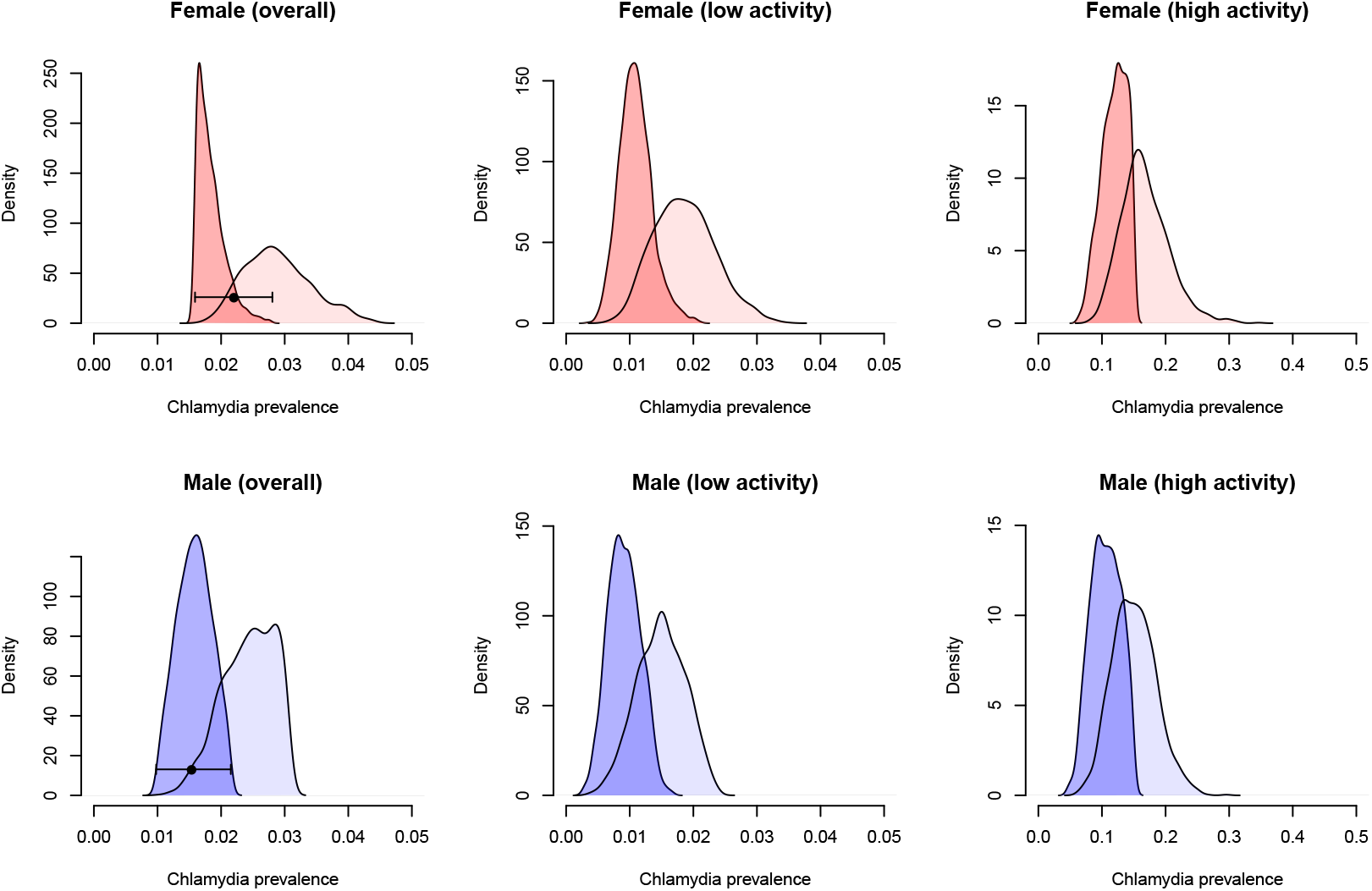
Posterior distributions of chlamydia prevalence (baseline scenario 1). Modelled prevalence in the presence and absence of interventions (asymptomatic treatment and partner notification) is shown as dark and light shaded areas. The black dot represents the prevalence for 16–34 year olds as estimated from Natsal-3 (together with 95% confidence intervals).

**Figure 4.**
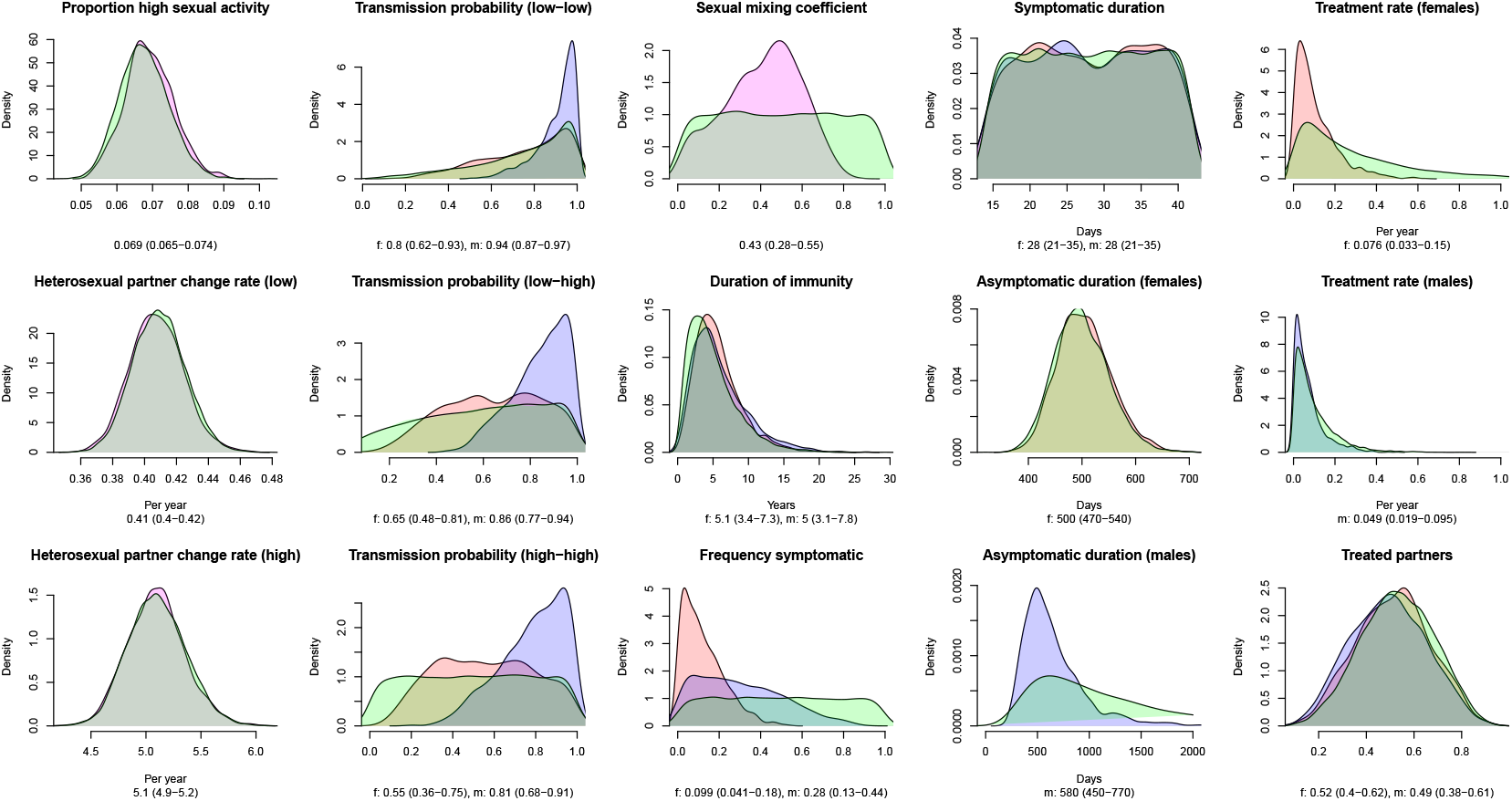
Prior and posterior distributions of model parameters (baseline scenario 1). Prior (green) and posterior distributions for females (red) and males (blue). The sexual mixing coefficient is the same for both sexes. Numbers below the horizontal axes represent the median and interquartile range.

### Who-Notifies-Whom (WNW) matrix

The PN module allowed us to construct the WNM matrix for female and male index cases (Tables 2 and 3). This matrix provides the chlamydia positivity in partners of index cases, stratified by the sexual activity group and the infection status of both index case and partner. Furthermore, we calculated the proportion of PNs that were initiated and the proportion of positive partners that were notified by each index case. For example, the overall chlamydia positivity in a male sexual partner of a female symptomatic index case of the low sexual activity group is 90% (IQR: 88%–93%) (most left column in Table 2). Most sexual partners of these index cases are asymptomatic men of the high sexual activity group (71%, IQR: 68%–74%). Symptomatically infected women of the low sexual activity group notify 28% (IQR: 16%–40%) of all male sexual partners. Due to the high chlamydia positivity in these partners, this index-partner combination contributes to 42% (IQR: 29%–51%) of all PNs that identify a chlamydia-positive partner. In summary, we can show that chlamydia positivity is highest in sexual partners of symptomatic index cases that belong to the low sexual activity group. The notified partners are typically asymptomatic and belong to the high sexual activity group, i.e., are those infected individuals that will contribute most to onward transmission. This general pattern is even more accentuated if index cases are men (3).

**Table 2.**
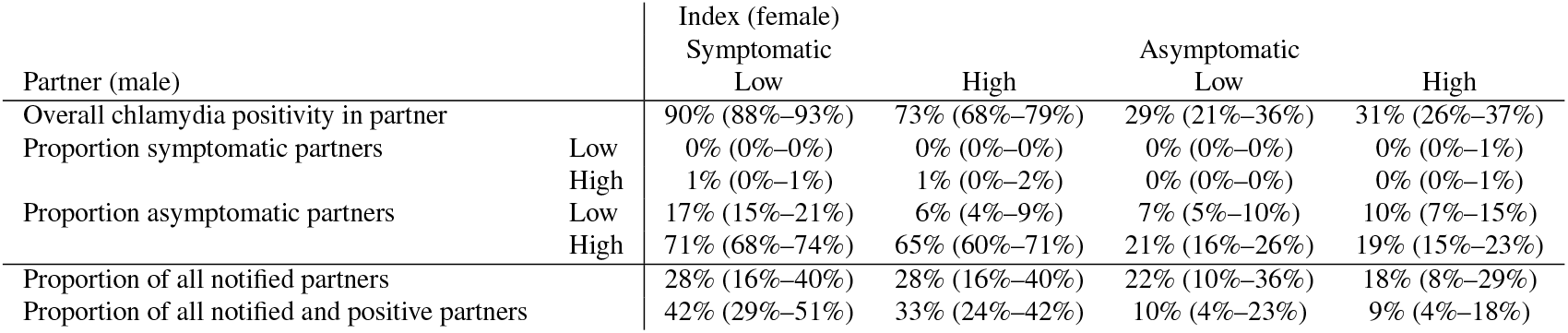
Who-Notifies-Whom (WNW) matrix for female index cases (scenario 1). Example: The overall chlamydia positivity in a male sexual partner of a female symptomatic index case of the low sexual activity group is 90%. The same index cases notify 28% of all male sexual partners, contributing to 42% of all notified and positive partners. Numbers are given as median and interquartile range.

**Table 3.** Who-Notifies-Whom (WNW) matrix for male index cases (scenario 1). Example: The overall chlamydia positivity in a female sexual partner of a male symptomatic index case of the low sexual activity group is 91%. The same index cases notify 40% of all female sexual partners, contributing to 48% of all notified and positive partners. Numbers are given as median and interquartile range.

| Partner (female) |  | Index (male) |  | Asymptomatic |  |
| --- | --- | --- | --- | --- | --- |
|  |  | Symptomatic |  | Low | High |
|  |  | Low | High |  |  |
| Overall chlamydia positivity in partner |  | 91% (88%–93%) | 79% (75%–83%) | 20% (14%–29%) | 47% (41%–53%) |
| Proportion symptomatic partners | Low | 0% (0%–0%) | 0% (0%–0%) | 0% (0%–0%) | 0% (0%–0%) |
|  | High | 0% (0%–1%) | 0% (0%–1%) | 0% (0%–0%) | 0% (0%–0%) |
| Proportion asymptomatic partners | Low | 20% (17%–24%) | 9% (6%–12%) | 7% (6%–9%) | 18% (14%–23%) |
|  | High | 70% (66%–73%) | 69% (63%–74%) | 13% (8%–21%) | 28% (24%–32%) |
| Proportion of all notified partners |  | 40% (31%–48%) | 41% (30%–49%) | 7% (2%–18%) | 6% (2%–15%) |
| Proportion of all notified and positive partners |  | 48% (40%–54%) | 42% (34%–49%) | 2% (0%–6%) | 4% (1%–10%) |

### APT intervention

Simulating the effects of APT on chlamydia transmission illustrates that it can be an effective method to reduce prevalence at the population-level (Fig 5). Increasing the numbers of treated partners from current levels in Britain (0.51, 95% CrI: 0.21–0.80, see Table 1) can result in a considerable reduction in prevalence after 5 years. For example, an increase of 25% would reduce chlamydia prevalence by 18% (95% CrI: 5%-44%) in both women and men. The relative reduction is slightly higher in women than men, and in high sexual activity individuals compared to low sexual activity individuals. The results show that the effects of APT on chlamydia transmission mainly stem from increasing the numbers of treated partners, as reducing the time to partner treatment from current levels (3.2 d, see Table 1) had only minor effects on chlamydia prevalence. Compared to the baseline scenario (scenario 1), the effects of APT were somewhat lower in scenario 2 (Fig S6) but higher in scenario 3 (Fig S5).

**Figure 5.**
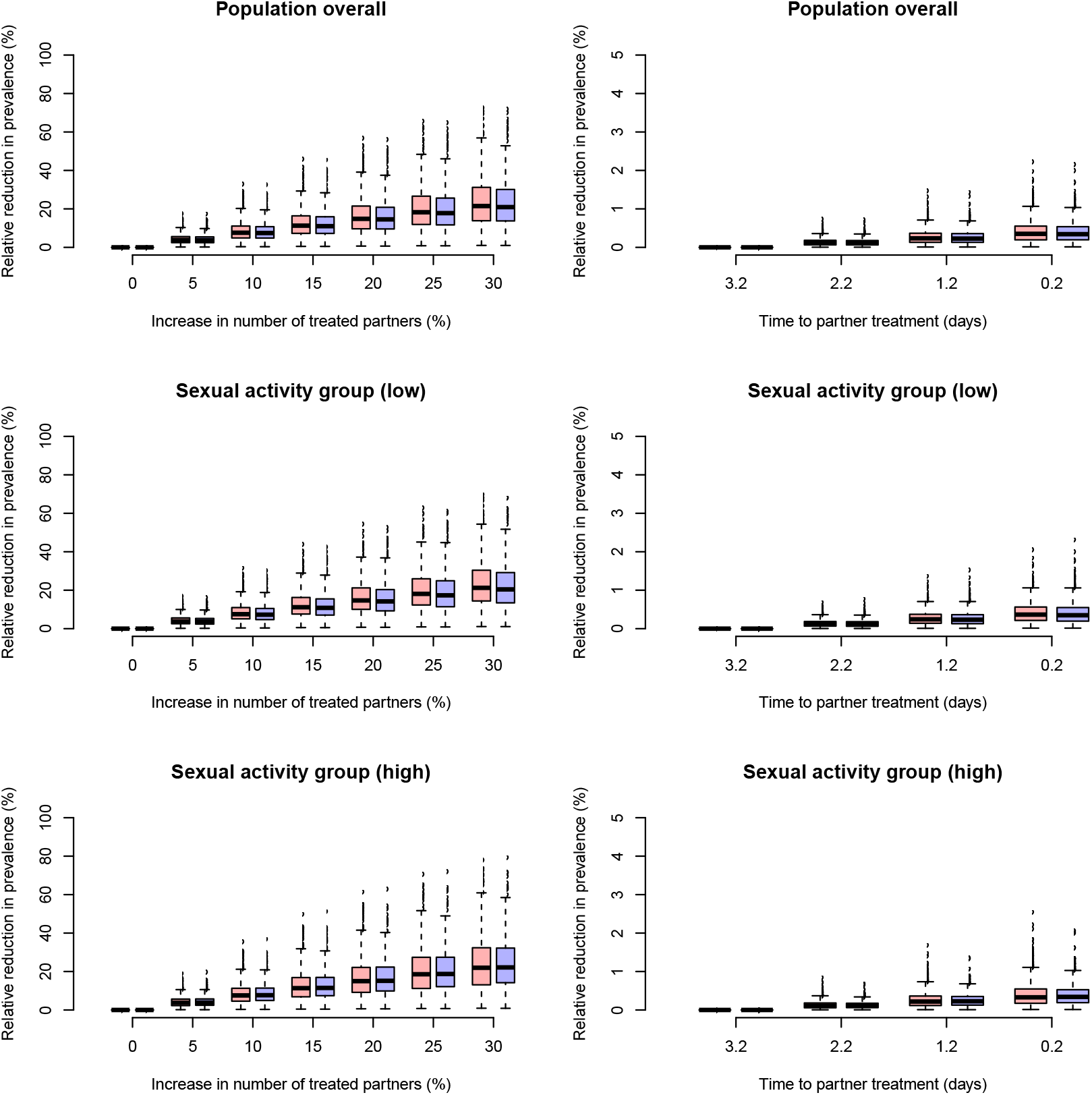
Projected effect of accelerated partner therapy (APT) on chlamydia prevalence after 5 years (baseline scenario 1). APT is modelled as an increase in the number of treated partners (left panels) or a reduction in the time to partner treatment (right panels). Changes in prevalence are given for females (red) and males (blue). Note the difference in scales of the axes between the left and right panels.

## DISCUSSION

Using a novel chlamydia transmission model, we explored the expected effects of APT on chlamydia prevalence compared to standard PN in Britain. We found that chlamydia positivity is highest in partners of symptomatic index cases with low sexual activity, whereas the infected partners are typically asymptomatic and highly sexually active. Conducting PN for this particular index-partner combination will thus be most effective for preventing further transmission. Increasing the number of treated partners from current levels by 25% would reduce chlamydia prevalence by 18% (95% CrI: 5%-44%) in both women and men within 5 years. In contrast, reducing the time to partner treatment alone had a minor effect on reducing prevalence. Together, these results suggest that PN typically identifies sexual partners that are likely to further transmit chlamydia, and that APT in particular has the potential to further reduce prevalence through an increase in PN uptake.

The main strength of our model is the PN module, which allowed us to construct a WNW matrix and look into the process of PN for chlamydia in great detail. In contrast to other modelling studies, we used the PN module to identify the index-partner combinations that result in the highest chlamydia positivity in partners and yield the greatest impact on reducing further transmission. Compared to other deterministic, population-based modelling approaches that limit PN to current sexual partners (Heijne et al., 2011; Rönn et al., 2019), our model allowed us to track the most recent partner for all index cases. Using a Bayesian modelling framework, we were also able to account for the considerable uncertainty in model parameters. Consideration of the specific distribution of chlamydia infections according to an individual’s sexual activity, has been suggested as an important aspect of STI transmission models (Althaus et al., 2012b). We therefore calibrated the model, not only to the current chlamydia prevalence in women and men, but also to chlamydia prevalence in different sexual activity groups and the expected chlamydia prevalence in the absence of asymptomatic testing and PN. The modelling framework can be further adapted for calibration to additional epidemiological data, such as chlamydia positivity in partners of index cases, and can therefore be tailored to the results of randomised controlled trials.

Our study has some limitations. First, we considered notification of the index case’s most recent partner only. This was a necessary simplification of our modelling framework. As the average number of notified partners is typically below one, we expect that including notification of additional partners in our model would not substantially affect our results. Second, we did not consider reinfection of index case cases by untreated partners. The modelled effects of APT on reducing chlamydia prevalence could thus be considered as best-case scenarios. Third, we did not consider different age groups and age-specific sexual mixing patterns (Smid et al., 2018), men having sex with men (MSM) or different ethnic groups in our model. As most chlamydia infections are found in the population of young opposite-sex partners, and because sexual mixing is arguably the most critical aspect for PN, we do not think that ignoring these additional complexities would substantially impact our results. Finally, there remains considerable uncertainty with respect to test uptake, chlamydia diagnoses (Chandra et al., 2017) and their impact on chlamydia prevalence (Lewis and White, 2018; Smid et al., 2019). We addressed this problem by choosing broad prior distributions for the treatment rate in our model.

The modelled chlamydia positivity in partners of index cases, which is typically not reported for modelling studies, is in good agreement with empirical data from Britain that show an overall positivity of 62% (Public Health England, 2016) and 69% (Public Health England, 2018). While these values correspond to the overall positivity, the WNW matrix highlights that chlamydia positivity is expected to be higher in partners of symptomatic index cases, but lower in partners of asymptomatic index cases. The precise contribution of PN on limiting the spread of chlamydia remains a matter of debate (Althaus et al., 2014; Rönn et al., 2017). In this study, and similar to the results from Rö nn et al. (2019), we found that PN in general can play an important role in reducing the burden of chlamydia.

We showed that APT could effectively reduce ongoing chlamydia transmission in Britan through an increase in the numbers of partners treated. Current levels of PN uptake (the number of contacts per index case who were reported as having attended a sexual health service within four working weeks of the date of the PN consultation) are below the standard of 0.6 (Public Health England, 2018) which suggests that methods that aim to increase uptake to this level and beyond should be considered for PN. Furthermore, it will be important to continue monitoring the levels of PN uptake over time. In contrast to increasing PN uptake, reducing the time to partner treatment appears to be less critical for increasing the effectiveness of PN. Average times for attending health services are already quite short (3.2 days, (Public Health England, 2016)), but APT might still offer substantial benefits in situations where there are longer delay periods. Our study highlights that PN for chlamydia is a highly effective intervention due to its targeted approach. With the WNW matrix, we can show that PN naturally identifies the most important partners of index cases: asymptomatically infected individuals from the high sexual activity group who thus remain infected for a long time period and have a high sexual partner change rate.

A detailed understanding of the effects of PN on chlamydia transmission is critical for implementing optimal control strategies. The presented modelling framework can be used to study the population-level effects of different PN interventions, such as APT, in considerable detail, and is flexible enough to be readily adapted to different populations and intervention scenarios. Our results suggest that APT, through an increase in the number of partners treated, would be an effective method to reduce ongoing chlamydia transmission in Britain.

## Data Availability

All data and R code files are available on GitHub: https://github.com/calthaus/chlamydia-pn.

## Acknowledgments

Calculations were performed on UBELIX (http://www.id.unibe.ch/hpc), the HPC cluster at the University of Bern. Natsal-3 is a collaboration between University College London, London School of Hygiene and Tropical Medicine, National Centre for Social Research, Public Health England, and the University of Manchester. We thank the study participants, the team of interviewers from NatCen Social Research, and operations and computing staff from NatCen Social Research. The Natsal-3 study was approved by the Oxfordshire Research Ethics Committee A (Ref: 10/H0604/27).

## Author contributions

All authors conceived and designed the study. CLA developed the mathematical model, performed model simulations, and wrote the manuscript. All authors contributed to the interpretation of the results, commented on the manuscript, and approved the final version of the manuscript.

## S1 SUPPORTING INFORMATION

### S1.1 Model calibration

#### S1.1.1 Chlamydia prevalence

**Figure S1.**
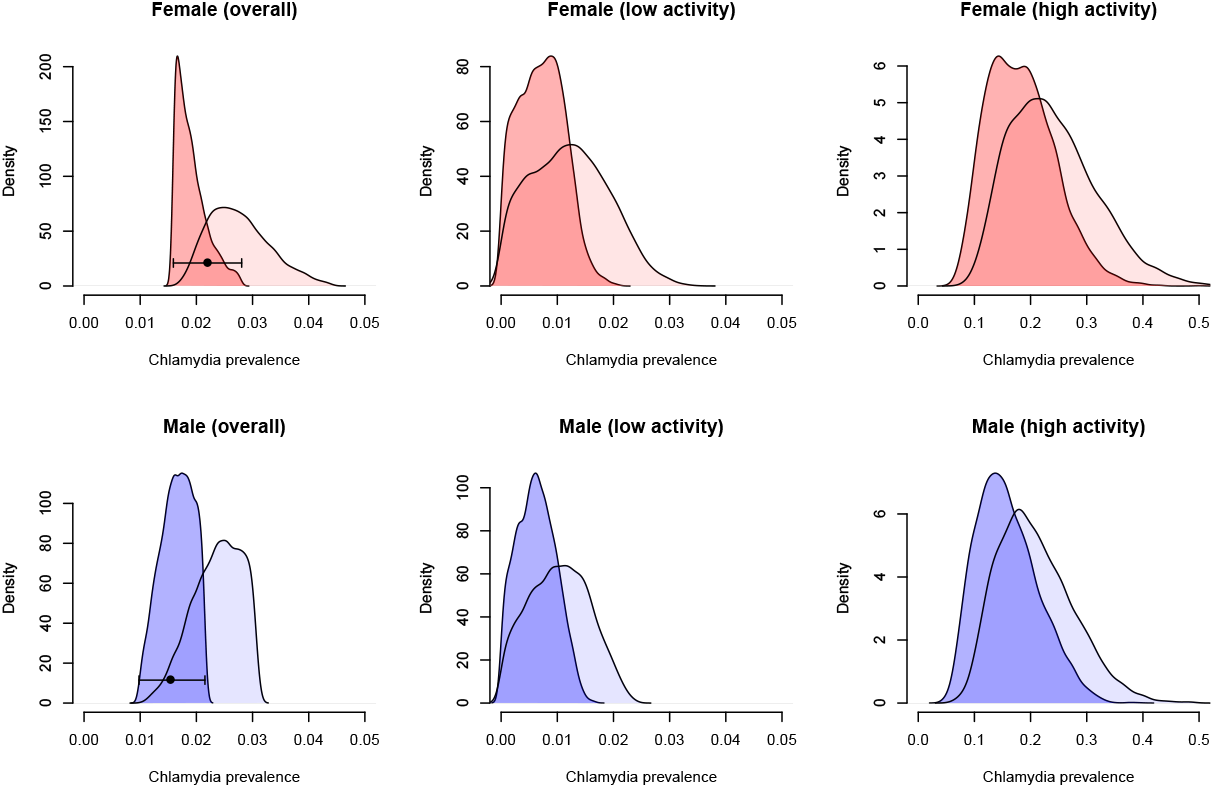
Posterior distributions of chlamydia prevalence (baseline scenario 2). Modelled prevalence in the presence and absence of interventions (asymptomatic treatment and partner notification) is shown as dark and light shaded areas. The black dot represents the prevalence for 16–34 year olds as estimated from Natsal-3 (together with 95% confidence intervals).

**Figure S2.**
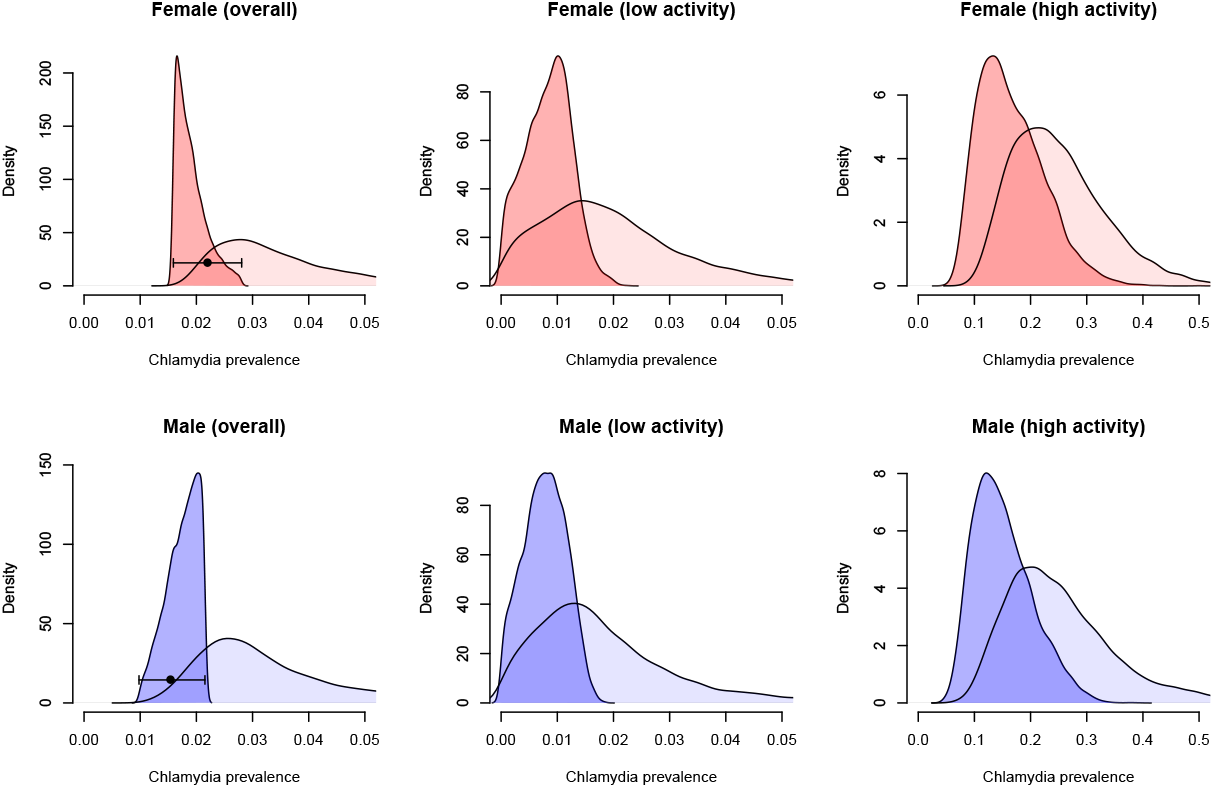
Posterior distributions of chlamydia prevalence (baseline scenario 3). Modelled prevalence in the presence and absence of interventions (asymptomatic treatment and partner notification) is shown as dark and light shaded areas. The black dot represents the prevalence for 16–34 year olds as estimated from Natsal-3 (together with 95% confidence intervals).

#### S1.1.2 Posterior distributions

**Figure S3.**
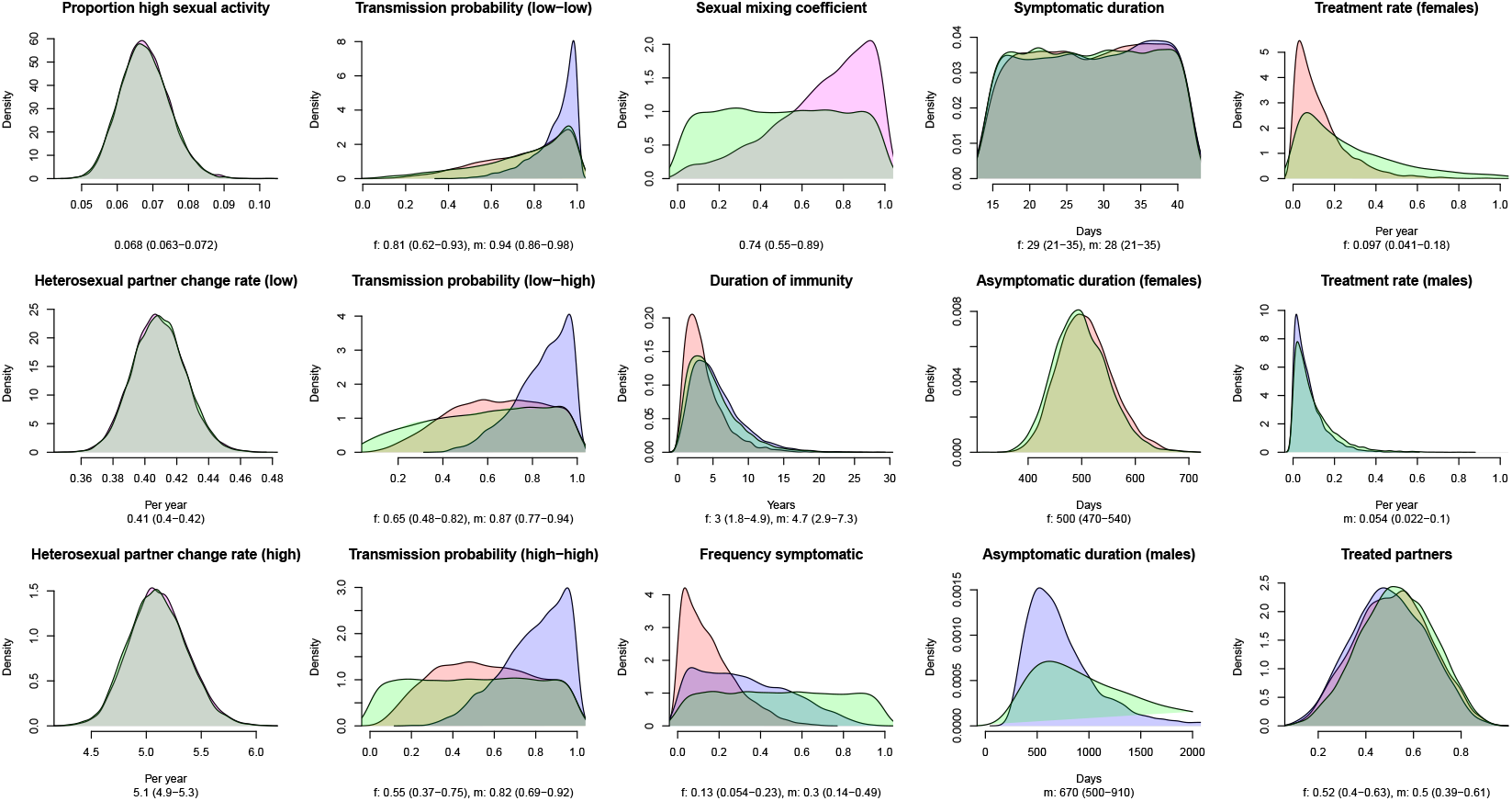
Prior and posterior distributions of model parameters (baseline scenario 2). Prior (green) and posterior distributions for females (red) and males (blue). The sexual mixing coefficient is the same for both sexes. Numbers below the horizontal axes represent the median and interquartile range.

**Figure S4.**
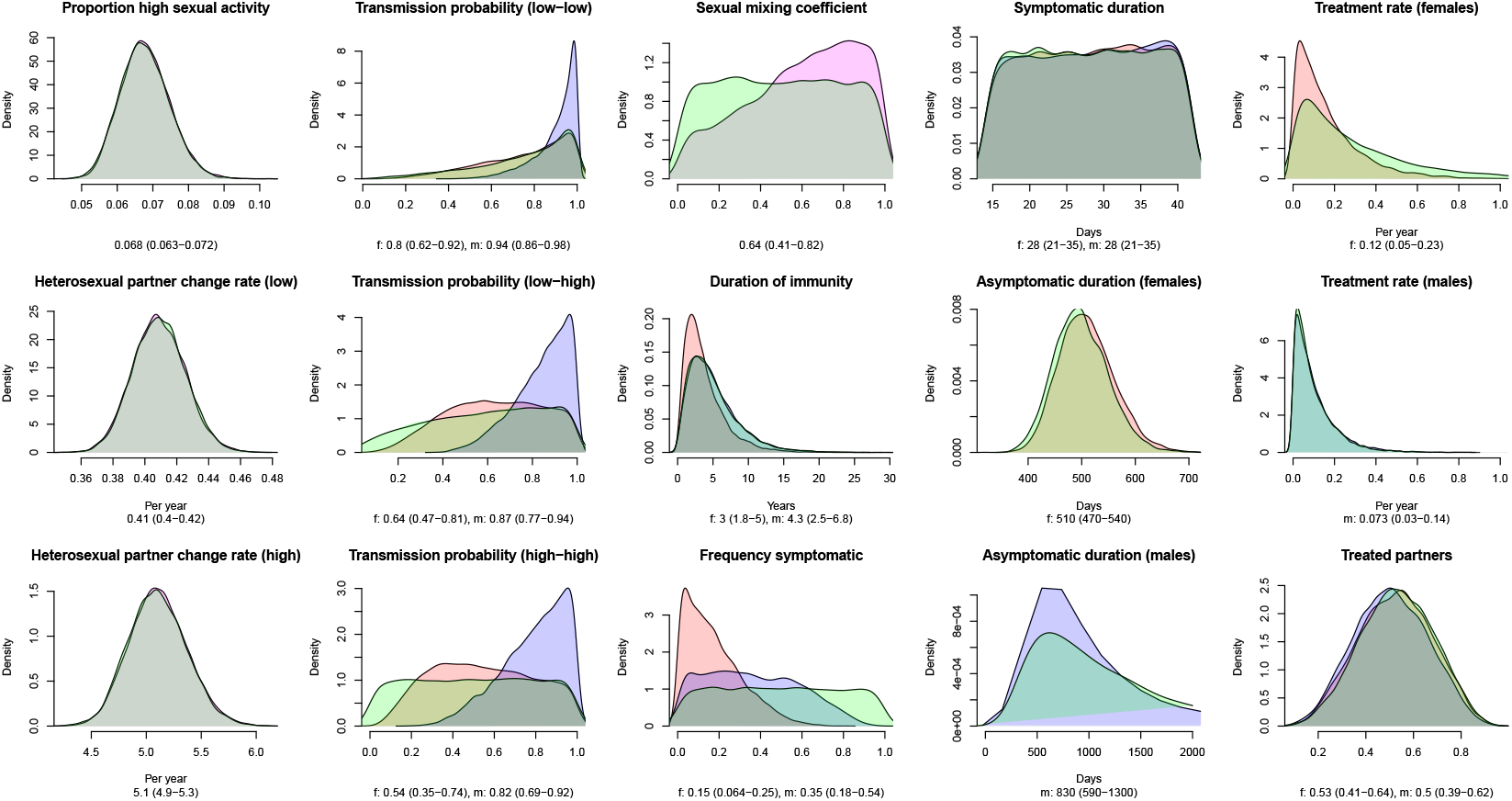
Prior and posterior distributions of model parameters (baseline scenario 3). Prior (green) and posterior distributions for females (red) and males (blue). The sexual mixing coefficient is the same for both sexes. Numbers below the horizontal axes represent the median and interquartile range.

#### S1.1.3 Who-Notifies-Whom (WNW) matrix

**Table S1.** Who-Notifies-Whom (WNW) matrix for female index cases (scenario 3). Numbers are given as median and interquartile range.

| Partner (male) |  | Index (female) |  | Asymptomatic |  |
| --- | --- | --- | --- | --- | --- |
|  |  | Symptomatic |  | Low | High |
|  |  | Low | High | Low | High |
| Uninfected |  | NA | NA | NA | NA |
| Symptomatic | Low | 0% (0%–0%) | 0% (0%–0%) | 0% (0%–0%) | 0% (0%–1%) |
|  | High | 1% (0%–2%) | 1% (0%–2%) | 0% (0%–0%) | 0% (0%–1%) |
| Asymptomatic | Low | 20% (16%–25%) | 3% (1%–6%) | 10% (7%–14%) | 6% (3%–10%) |
|  | High | 68% (64%–72%) | 68% (62%–74%) | 24% (18%–31%) | 23% (18%–28%) |
| Chlamydia positivity in partner |  | 91% (88%–93%) | 74% (68%–79%) | 35% (27%–45%) | 31% (26%–36%) |
| Proportion of notified partners |  | 18% (9%–31%) | 37% (20%–52%) | 13% (5%–27%) | 21% (9%–36%) |
| Proportion positive among notified |  | 29% (15%–43%) | 42% (27%–56%) | 7% (2%–20%) | 11% (4%–22%) |

**Table S2.** Who-Notifies-Whom (WNW) matrix for male index cases (scenario 3). Numbers are given as median and interquartile range.

| Partner (female) |  | Index (male) |  | Asymptomatic |  |
| --- | --- | --- | --- | --- | --- |
|  |  | Symptomatic |  | Low | High |
|  |  | Low | High | Low | High |
| Uninfected |  | NA | NA | NA | NA |
| Symptomatic | Low | 0% (0%–0%) | 0% (0%–0%) | 0% (0%–0%) | 0% (0%–0%) |
|  | High | 0% (0%–1%) | 0% (0%–1%) | 0% (0%–0%) | 0% (0%–1%) |
| Asymptomatic | Low | 21% (18%–25%) | 4% (2%–9%) | 6% (5%–8%) | 11% (5%–18%) |
|  | High | 67% (63%–71%) | 71% (65%–76%) | 8% (3%–14%) | 32% (26%–38%) |
| Chlamydia positivity in partner |  | 90% (87%–92%) | 78% (73%–82%) | 15% (9%–22%) | 45% (40%–53%) |
| Proportion of notified partners |  | 27% (16%–39%) | 48% (34%–63%) | 6% (2%–16%) | 8% (3%–20%) |
| Proportion positive among notified |  | 35% (21%–47%) | 50% (38%–64%) | 1% (0%–4%) | 5% (2%–14%) |

**Table S3.**
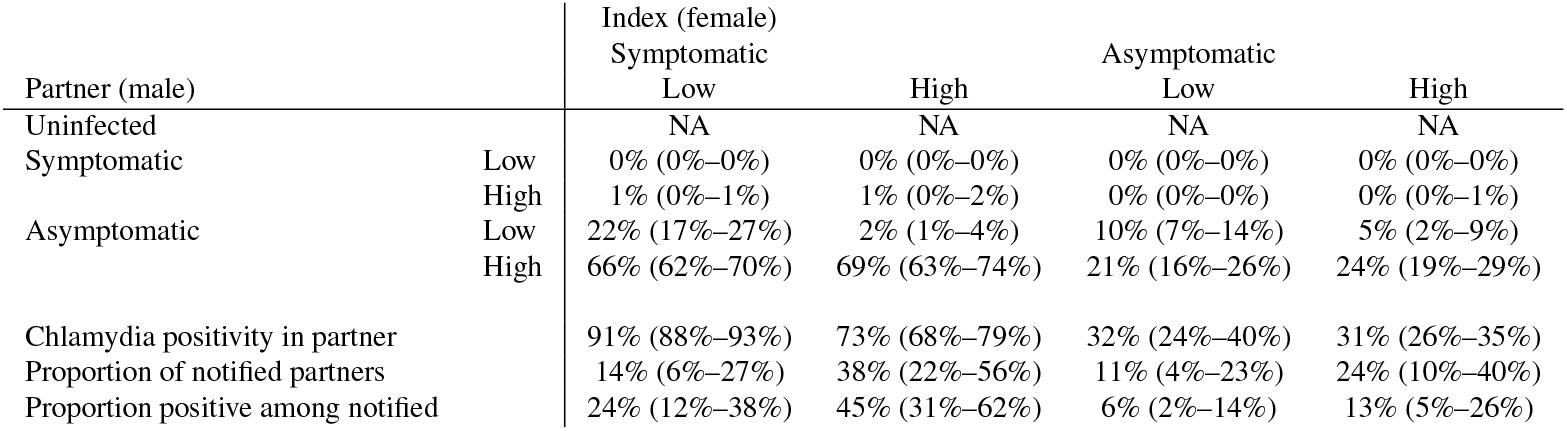
Who-Notifies-Whom (WNW) matrix for female index cases (scenario 2). Numbers are given as median and interquartile range.

**Table S4.**
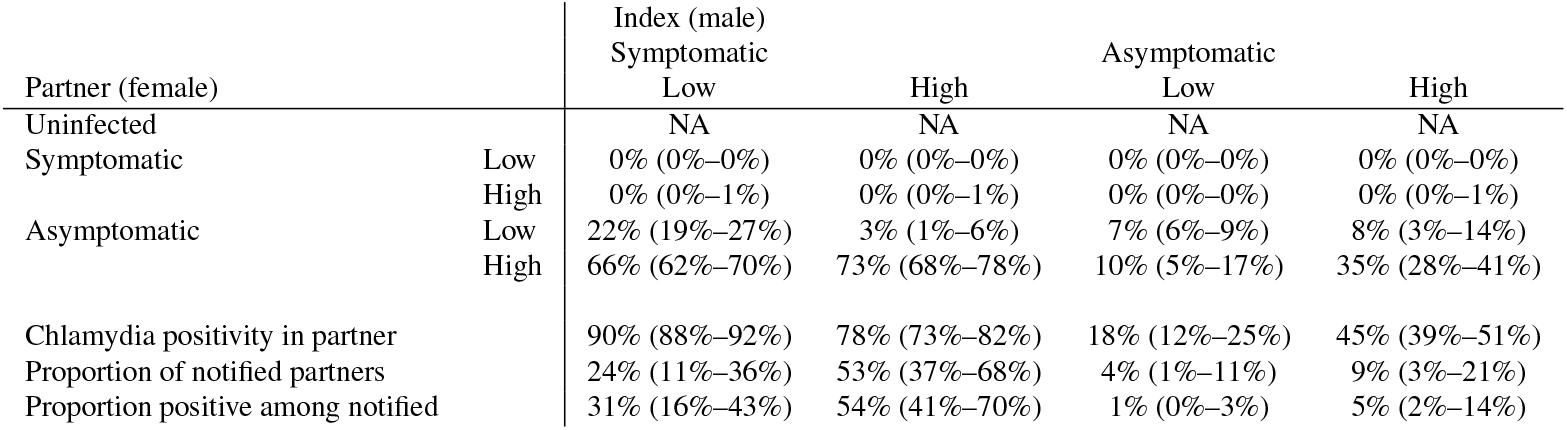
Who-Notifies-Whom (WNW) matrix for male index cases (scenario 2). Numbers are given as median and interquartile range.

#### S1.1.4 Effect of accelerated partner therapy (APT)

**Figure S5.**
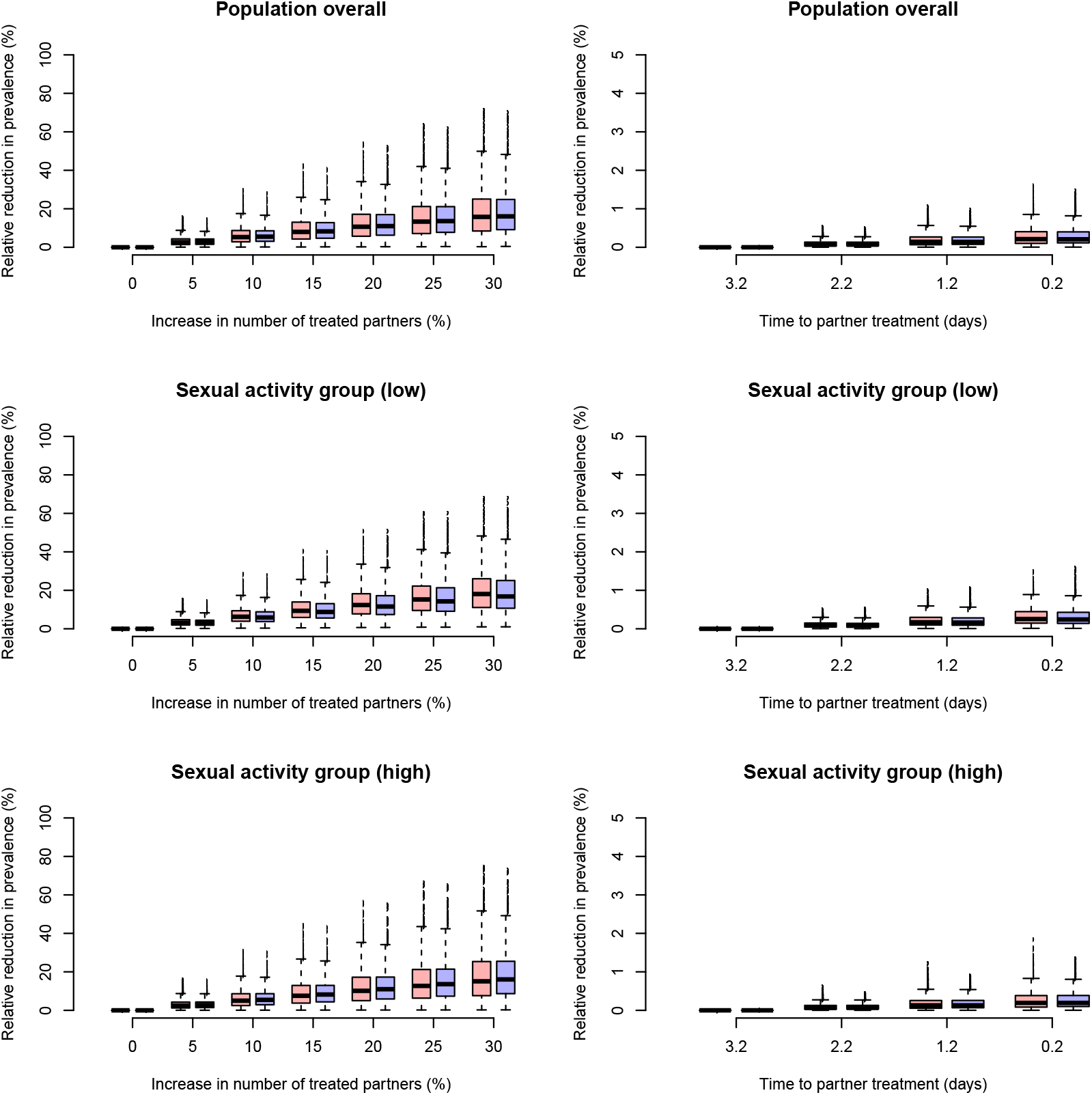
Projected effect of accelerated partner therapy (APT) on chlamydia prevalence after 5 years (scenario 2). APT is modelled as an increase in the number of treated partners (left panels) or a reduction in the time to partner treatment (right panels). Changes in prevalence are given for females (red) and males (blue). Note the difference in scales of the axes between the left and right panels.

**Figure S6.**
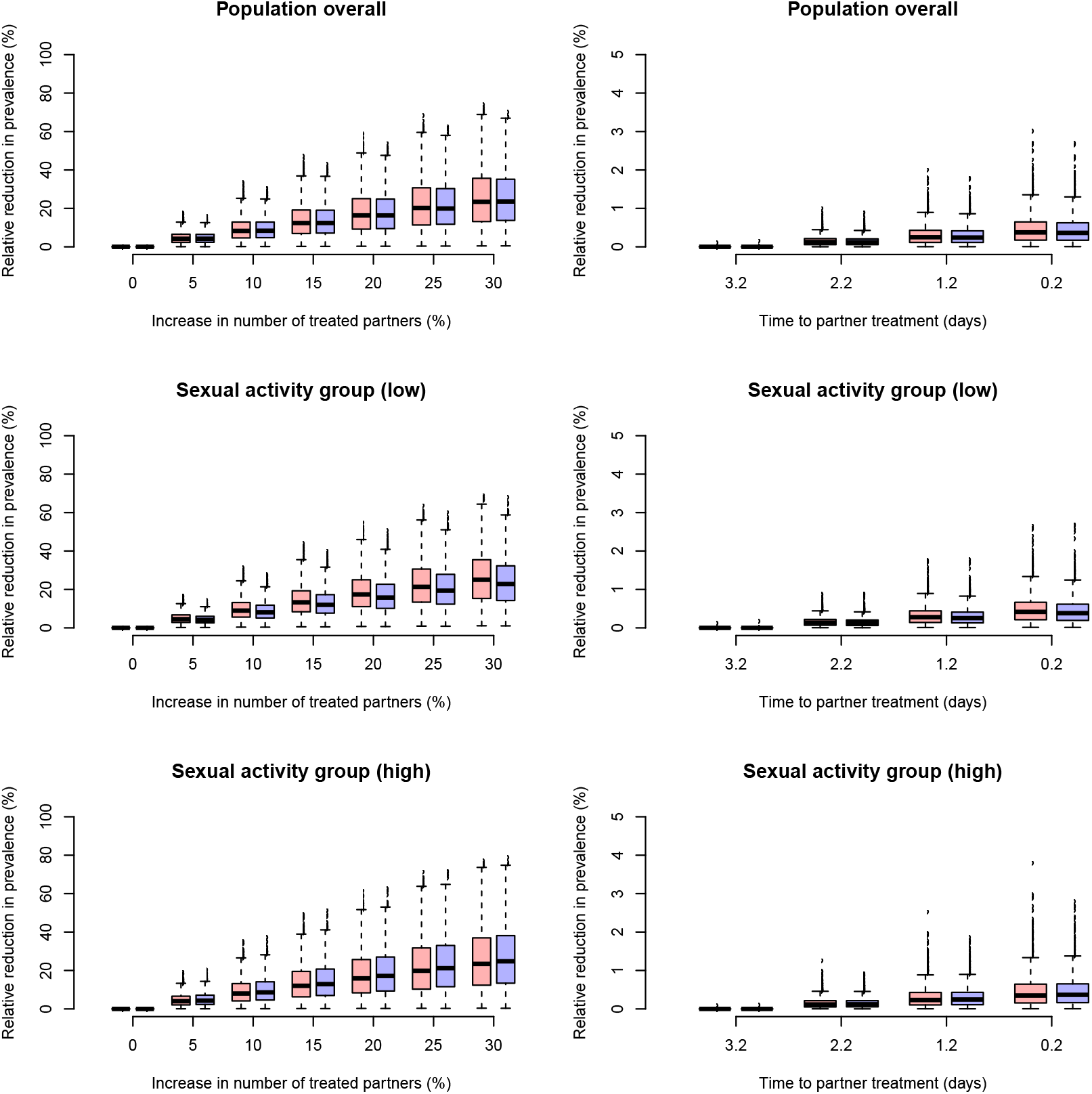
Projected effect of accelerated partner therapy (APT) on chlamydia prevalence after 5 years (scenario 3). APT is modelled as an increase in the number of treated partners (left panels) or a reduction in the time to partner treatment (right panels). Changes in prevalence are given for females (red) and males (blue). Note the difference in scales of the axes between the left and right panels.

